# Modelling of Systemic versus Pulmonary Chloroquine Exposure in Man for COVID-19 Dose Selection

**DOI:** 10.1101/2020.04.24.20078741

**Authors:** Ghaith Aljayyoussi, Rajith KR Rajoli, Henry Pertinez, Shaun H Pennington, W. David Hong, Paul M. O’Neill, Andrew Owen, Steve A Ward, Giancarlo A Biagini

**Author notes:** Corresponding Author: **address:** Centre for Drugs & Diagnostics, Department of Tropical Disease Biology, Liverpool School of Tropical Medicine, Pembroke Place, Liverpool L3 5QA, UK, **email:**. **Funding:** Medical Research Council, MR/S020411/1. All authors have confirmed no conflict of interest.

## Abstract

Chloroquine has attracted intense attention as a potential clinical candidate for prevention and treatment of COVID-19 based on reports of *in-vitro* efficacy against SARS-CoV-2. While the pharmacokinetic-pharmacodynamic (PK-PD) relationship of chloroquine is well established for malaria, there is sparse information regarding its dose-effect relationship in the context of COVID-19.

Here, we explore the PK-PD relationship of chloroquine for COVID-19 by modelling both achievable systemic and pulmonary drug concentrations. Our data indicate that the standard anti-malarial treatment dose of 25mg/kg over three days does not deliver sufficient systemic drug exposures for the inhibition of viral replication. In contrast, PK predictions of chloroquine in the lungs using *in-vivo* data or human physiologically-based PK models, suggest that doses as low as 3mg/kg/day for 3 days could deliver exposures that are significantly higher than reported antiviral-EC_90_s for up to a week. Moreover, if pulmonary exposure is a driver for prevention, simulations show that chronic daily dosing of chloroquine may be unnecessary for prophylaxis purposes. Instead, once weekly doses of 5mg/kg would be sufficient to achieve a continuous cover of therapeutically active pulmonary exposures.

These findings reveal a highly compartmentalised distribution of chloroquine in man that may significantly affect its therapeutic potential against COVID-19. The systemic circulation is shown as one site where chloroquine exposure is insufficient to inhibit SARS-CoV-2 replication. However, if therapeutic activity is driven by pulmonary exposure, it should be possible to reduce the chloroquine dose to safe levels. Carefully designed randomized controlled trials are urgently required to address these outstanding issues.

## INTRODUCTION

COVID-19 was declared as a global pandemic by the World Health Organisation (WHO) on 12^th^ of March 2020 (1). At the time of writing, there have been 2.7 million confirmed cases of disease and more than 190,000 deaths (2). Currently, there is no standard chemotherapeutic agent recommended for treatment or prevention.

The 4-aminoquinoline anti-malarial drug, chloroquine (CQ), has shown promising *in-vitro* activity against cultured SARS-CoV-2 with a comparable potency to anti-viral drugs, including remdesivir (CQ EC_50_ = 1.13µM, remdesivir EC_50_ = 0.77µM) (3). Limited trials have also demonstrated potential activity of the drug in patients with COVID-19 related pneumonia (4, 5). Currently a large clinical trial is taking place, where thousands of health workers will receive CQ or hydroxychloroquine (HCQ) to assess the two drugs’ prophylactic potential (6). CQ has also shown some pre-clinical anti-viral activity against a number of other intracellular pathogens including the closely related SARS-COV virus, albeit with 8-fold less potency than is currently reported for SARS-CoV-2 (7).

CQ was first developed for the treatment of malaria in the mid 1940s (8). It has since been deployed globally (many million human doses) for the treatment of malaria in populations across different continents and it continues to be used today in areas where malaria is endemic and where low levels of resistance to the compound are observed (9). Despite its widespread use and good safety profile, CQ is still considered to have a narrow therapeutic window with potential cardiotoxicity (10). Crucially, the drug has exhibited significant signs of lethal toxicity in its application (with large doses) in COVID-19 patients (11). Warnings of the potential cardiac toxicity for its use in the treatment of COVID-19 have already been made (12). The risk of ventricular arrhythmias following QT interval prolongation is expected to have more detrimental effects in patients in special groups such as the elderly with underlying cardiac disease, who are also at highest risk from COVID-19 complications (13). Furthermore, the chronic use of drugs of this class in patients with rheumatoid arthritis (RA) has been linked to long-term retinal damage (14).

The clinical therapeutic potential of CQ for prevention or treatment of COVID-19 is currently unknown. Here, we used population PK-PD modelling to assess the ability of achieving efficacious systemic concentrations of drug at doses considered safe and compared these to optimal dosage regimens required to achieve the maximal therapeutic drug concentration within the lung, the target organ for COVID-19, using two different methods (for comparison and validation) to predict human lung exposures. Our PK-PD approach gives particular attention to local pulmonary concentrations of the drug in order to predict its potential efficacy at different dosing levels using a comprehensive PK-PD relationship derived from published *in-vitro data*. We show here that the standard anti-malarial dose of 25mg/kg over three days starting with 15mg/kg on the first day (15) is therapeutically inadequate if systemic exposure is the driver of efficacy. In contrast, predicted concentrations in the lung are significantly higher than reported *in-vitro* EC_50_ suggesting efficacy would be achieved with much lower doses. Our analyses of pulmonary exposures show that doses as low as 3mg/kg administered for 3 days would be potentially sufficient to exert the maximal anti-viral activity that is predicted for this compound at the level of the lung. This dose is comparable to the dose used in rheumatoid arthritis (RA) treatments, deemed to be generally safe in the RA population (16), at least in the short term (<600g CQ total cumulative dose (17)). Furthermore, we show that long term efficacious cover of the drug can be achieved by once weekly dosing rather than chronic dosing as currently planned if pulmonary exposure drives prophylactic efficacy (6).

## METHODS

### i. *in-vivo* based lung PK modelling and extrapolation to human

To simulate CQ drug concentration profiles in human plasma we used population pharmacokinetic (PK) parameters reported in healthy volunteers (18) where drug levels were measured in plasma. The parameters of the used model are shown in **Table S1**.

To estimate the clinical pulmonary exposure of CQ in human lungs, we extrapolated from experimental accumulation data for CQ within lungs of the rat as previously published (19). This was achieved by simultaneously fitting CQ plasma and lung exposures to predict parameters that describe drug partitioning to the lungs. The rat plasma was fitted with a 2 compartment 1^st^ order absorption model while the lung was fitted as an open-loop configuration tissue compartment driven by the plasma concentration as a forcing function, with input/output distribution parameters expressed with the terms ***Q***_***lung_in***_ and ***Q***_***lung_out***_ (**Figure 1**).

**Figure 1.**
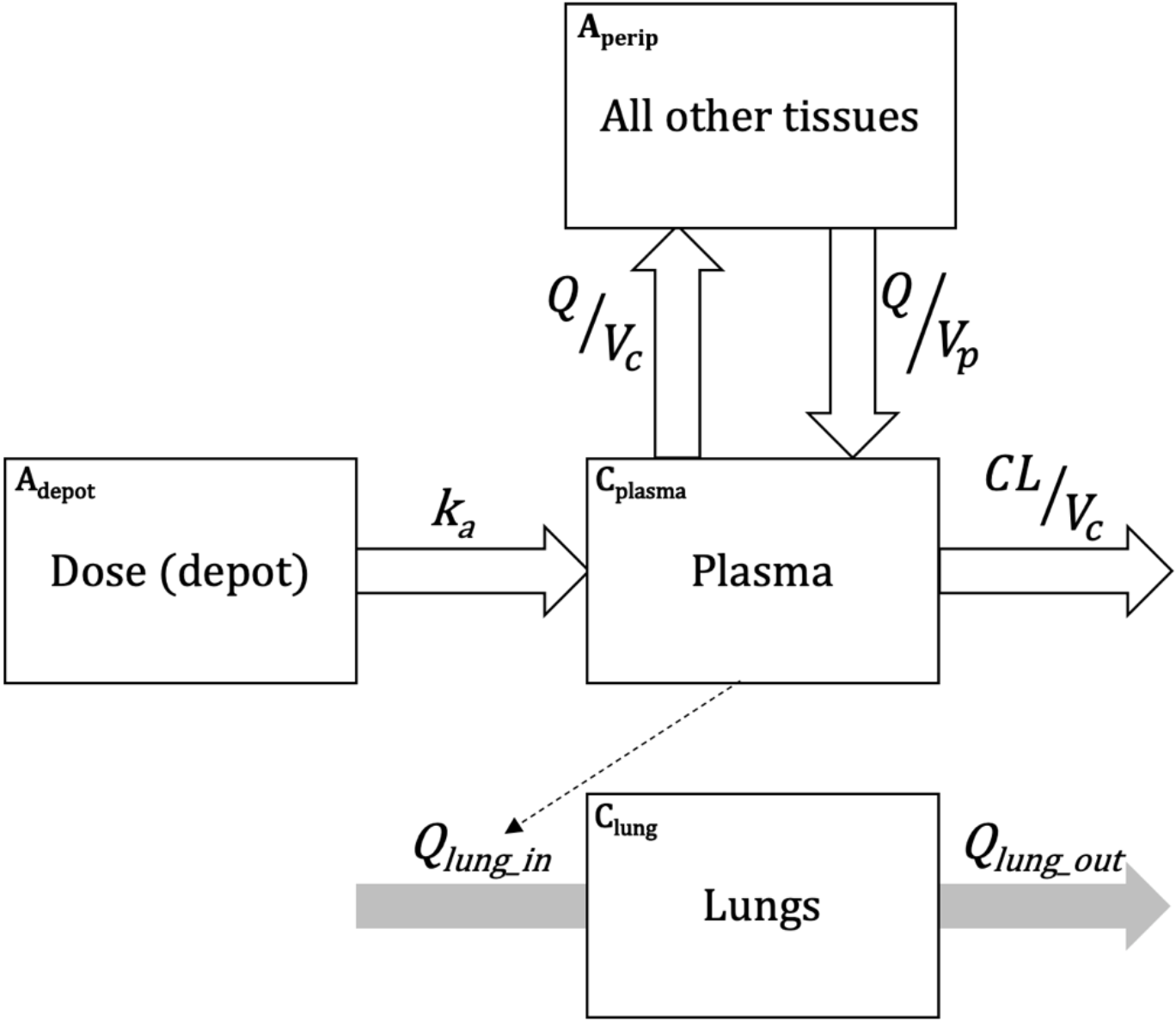
Schematic displaying the PK distribution model that was used to simulate CQ exposure in plasma and lungs of patients receiving it. Plasma exposure is based on parameters reported in humans while lung exposure parameters are derived from rat data using the same structural model.

The lung distribution parameters were then linked to the known population PK profile of CQ as previously published by using **Equations 1-4** (**Figure 1**).

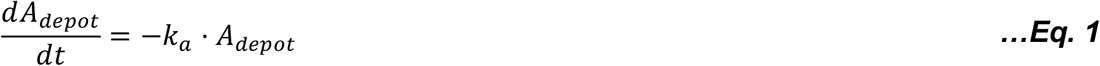

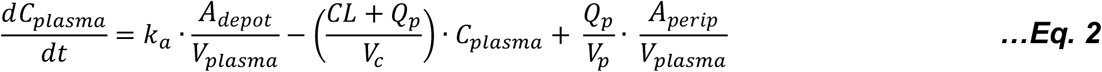

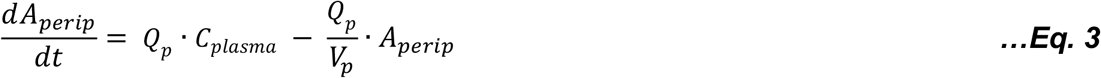

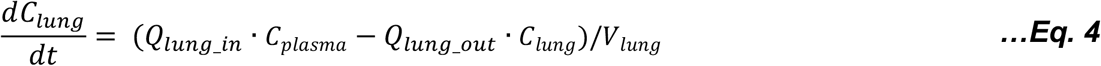

Where **A**_**depot**_ and **A**_**perip**_ represent drug mass in the gut and peripheral compartment and **C**_**plasma**_ and **C**_**lung**_ the concentrations in plasma and lung, respectively. **k**_**a**_ is the absorption rate constant from the dosing site depot to the central plasma compartment, **CL** the plasma clearance, **Q**_**p**_ is the intercompartmental clearance between the central and peripheral compartments. **V**_**plasma**_ (scaling volume of distribution for plasma) was estimated as part of the model fitting, and apparent (**V**_**lung**_) was fixed to a physiological value of 0.0021 L/kg [27], assuming rat weight of 0.25kg.

***Q***_***lung_in***_ and ***Q***_***lung_out***_ express the rate of drug accumulation in the lung as a function of its levels in the central plasma compartment while not directly linking it to the mass balance of that system, maintaining mass balance within the 2-compartment disposition model PK parameters (open-loop configuration). An estimate of the flow of drug into the lung is therefore partitioned out from the overall mass balance by the use of the two **Q** parameters.

Human lung exposure was therefore predicted using model equations 1-4, with human values for the 2-compartment plasma PK model (**k**_**a**_, **CL, Q**_**p**_ and **V**_**plasma**_) from Karunajeewa et al. (providing the plasma forcing function). **Q**_**lung_in**_ and **Q**_**lung_out**_ were scaled from rat to human assuming the ratio of **V**_**lung**_ to **V**_**plasma**_ was the same in both species respectively as follows:

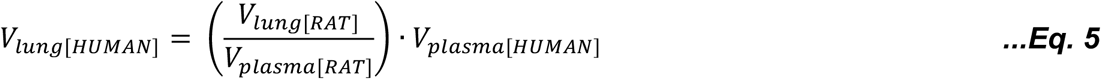

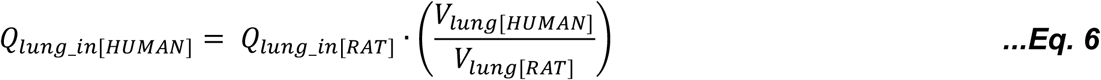

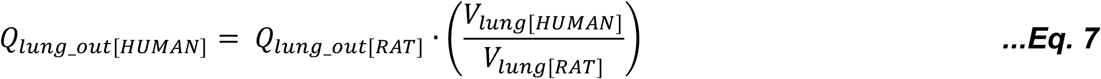

Model fitting/parameter estimation was performed using Monolix v. 2018R2 [26] operated via the IQR Tools package in R (v 3.5.3) (20, 21) and a summary of estimated parameters and %RSEs of estimates is provided in **Table S2**.

### ii. *in-vitro* based PBPK Modelling and human lung prediction

CQ plasma and lung concentrations were also simulated using a whole-body physiologically based PK (PBPK) model implemented in Matlab (Mathworks, v. 2019a). Rat and human PBPK models were designed. One hundred virtual healthy rats (weighing between 100-110g, similar to literature (22)) and humans (between 18-60 years and 40-120 kg) were simulated in this study. The model is blood-flow limited and assumes the distribution in organs and tissues is well-stirred, instant and uniform. For oral CQ, reabsorption from the colon was considered negligible.

The anatomy and physiology of both species were obtained from various literature sources (23-25). Drug disposition across various organs and tissues and tissue-to-plasma ratios were determined using published equations (26, 27). The drug specific parameters used in the human model are presented in **Table S3**.

Assuming the same protein binding as that of humans in rats, simulations using the rat PBPK model were compared to the observed data at a single 10 mg/kg dose similar to the literature (22). Lung concentrations over various time points were computed using the given tissue to plasma ratios and the plasma concentration from Adelusi et al. (22). Observed and simulated plasma and lung concentrations were compared, and the model was assumed to perform well if the mean simulated value was within 2-fold from the mean observed value (28).

After model verification in rats, the human PBPK model was also validated against oral data for a single 300 mg CQ dose (29). Subsequently, simulations in humans were performed with a single 5 mg/kg CQ dose and 5 mg/kg CQ weekly dose for 4 weeks to inform the plasma and lung concentrations.

### iii. Pharmacodynamic modelling

To estimate drug efficacy upon the SARS-CoV-2 virus clinically, the predicted pulmonary exposure estimated from the PK model was linked to anti-viral activity as derived from data published by Wang et al. (3) of the drug’s activity *in-vitro*. The data reported was used to generate an ***EC***_***50***_ value (concentration required to achieve the 50% of the drug’s maximum anti-viral-activity) and a ***hill*** constant (which expresses the steepness of the exposure-response curve). These parameters were used in ***Equation 8*** to link pulmonary exposure (expressed as ***C*** in the equation) to anti-viral activity.

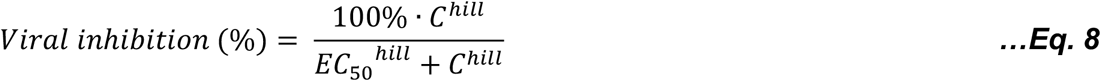

The pharmacodynamic values used in this simulation are shown in **Table S4**.

### iv. Monte-Carlo Simulations

Different dosing regimens of CQ, ranging from 0.25/mg/kg/day to 15mg/kg/day with 0.25mg/kg dose increments were sequentially simulated. We also used the inter-individual variability reported in Karunajeewa et al. Population PK analysis (18) for the 2-compartment model PK parameters and added an estimated 30% variation on lung partitioning parameters ***Q***_***lung_in***_ and ***Q***_***lung_out***_. For each dosing level, 1000 patients were simulated to estimate the percentage of patients who achieve a defined clinical target. All simulations were performed using IQRtools package for systems pharmacology and pharmacometrics version 1.1.1 through R version 3.5.3.

## RESULTS

### i. Quantification of CQ lung accumulation rate from rat data

**Figure 2** shows the fits generated from rat data reported in Adelusi et al. (19). The data shows a slow and continuous accumulation with a high extent of accumulation within the lung.

**Figure 2.**
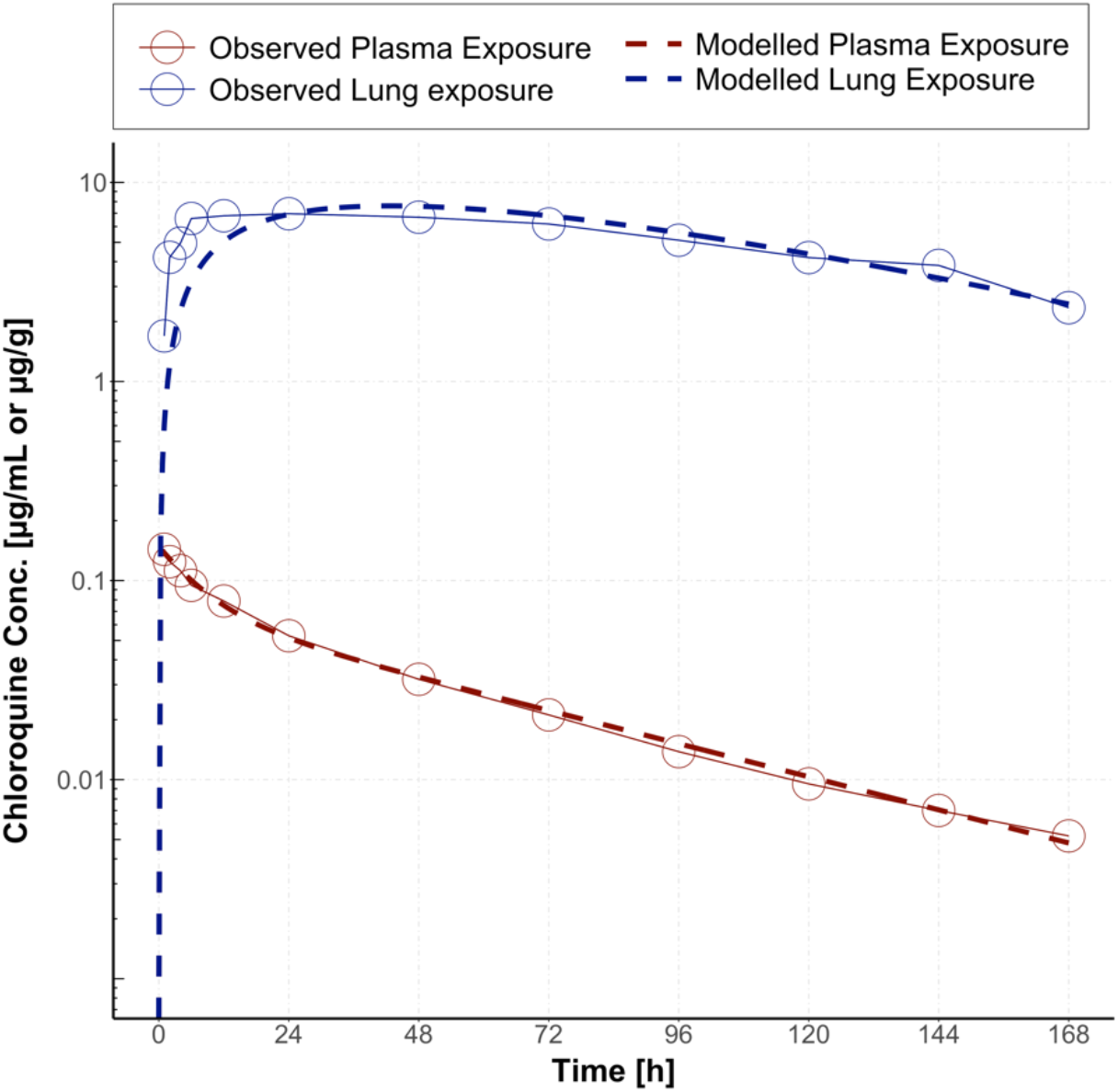
Modelling of rat plasma and lung exposure data. Red points represent reported plasma exposure, blue points reported lung exposure over time and the dashed lines represent the respected predicted exposure as predicting from fitting the rat data.

The PK parameters were estimated in the rat as per **Equations 1-4**. Exchange flow to and from the lung (***Q***_***lung_in***_ and ***Q***_***lung_out***_) were 0.394 L/h and 1.84 · 10^−3^ L/h respectively (All estimated PK parameters and %RSEs of estimates are in **Table S2**). The high lung:plasma ratio for CQ modelled here was confirmed by even higher ratios reported in other studies for the same drug where > 100-fold lung:blood ratios of CQ were reported at 24h post dose in the rat (30). The data used for modelling lung:plasma ratios in the rat in this work are hence the most conservative in this context.

### ii. Quantification of CQ lung accumulation using PBPK

**Figure 3** shows the simulated plasma and lung concentrations in humans. Both simulations, whether based on *in-vivo* rat data or utilising PBPK modelling using physicochemical properties of the drug result in similar predictions of CQ levels in the lung using two hypothetical dosing scenarios of 5mg/kg as a single dose or administered weekly. The PBPK model was performed completely independently from the *in-vivo* based model and was used as a validation method for outcomes of the *in-vivo* based simulation. Both models result in a nearly identical C_max_ level within the lung and both models predicted some 100-fold accumulation within the lung compared to plasma.

**Figure 3.**
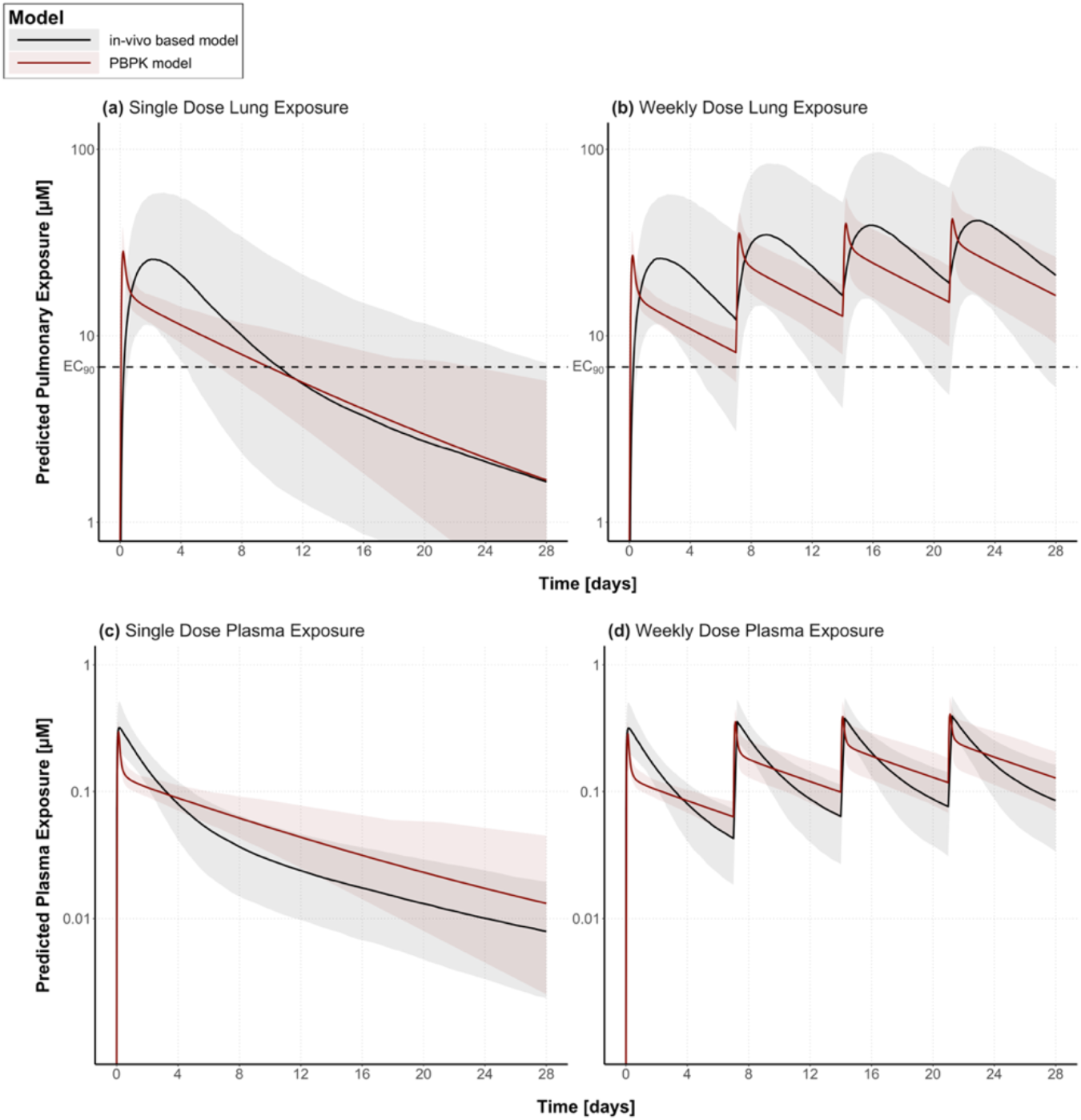
Predicted clinical exposure using the PBPK method (dark red line) or through using in-vivo rat extrapolation (black line). The predictions are for lung exposure (**a, b**) and plasma exposure (**c, d**). Shaded areas represent the 5-95 percentiles of each simulation. Comparison is shown for two dosing regimens: Single 5mg/kg dose (left) or a weekly 5mg/kg dose (right). For plasma exposure the in-vivo based model utilised population PK data published elsewhere, and the predicted plasma exposure (black line) is hence that observed in a clinical population, while for PBPK model the predicted exposure was generated using in-vitro data and PBPK modelling.

### iii. Predicting CQ’s therapeutic activity within the lungs

Dosing regimens ranging from (1mg/kg – 4mg/kg daily for 3 days) as well as the standard anti-malarial regimen (25mg/kg over 4 doses in three days) were simulated. These included the RA treatment daily dosing level (4mg/kg/day but for 3 days only rather than chronically) to estimate lung exposure and activity. The results show that when assuming that the lung exposure to drug is the driver for activity, lower doses of 3mg/kg/day given for three days would be sufficient to establish >90% of activity in the lung for up to a week after administration and that the anti-malarial standard dose of 25mg/kg over 3 days seems to be redundantly high (**Figure 4**).

**Figure 4.**
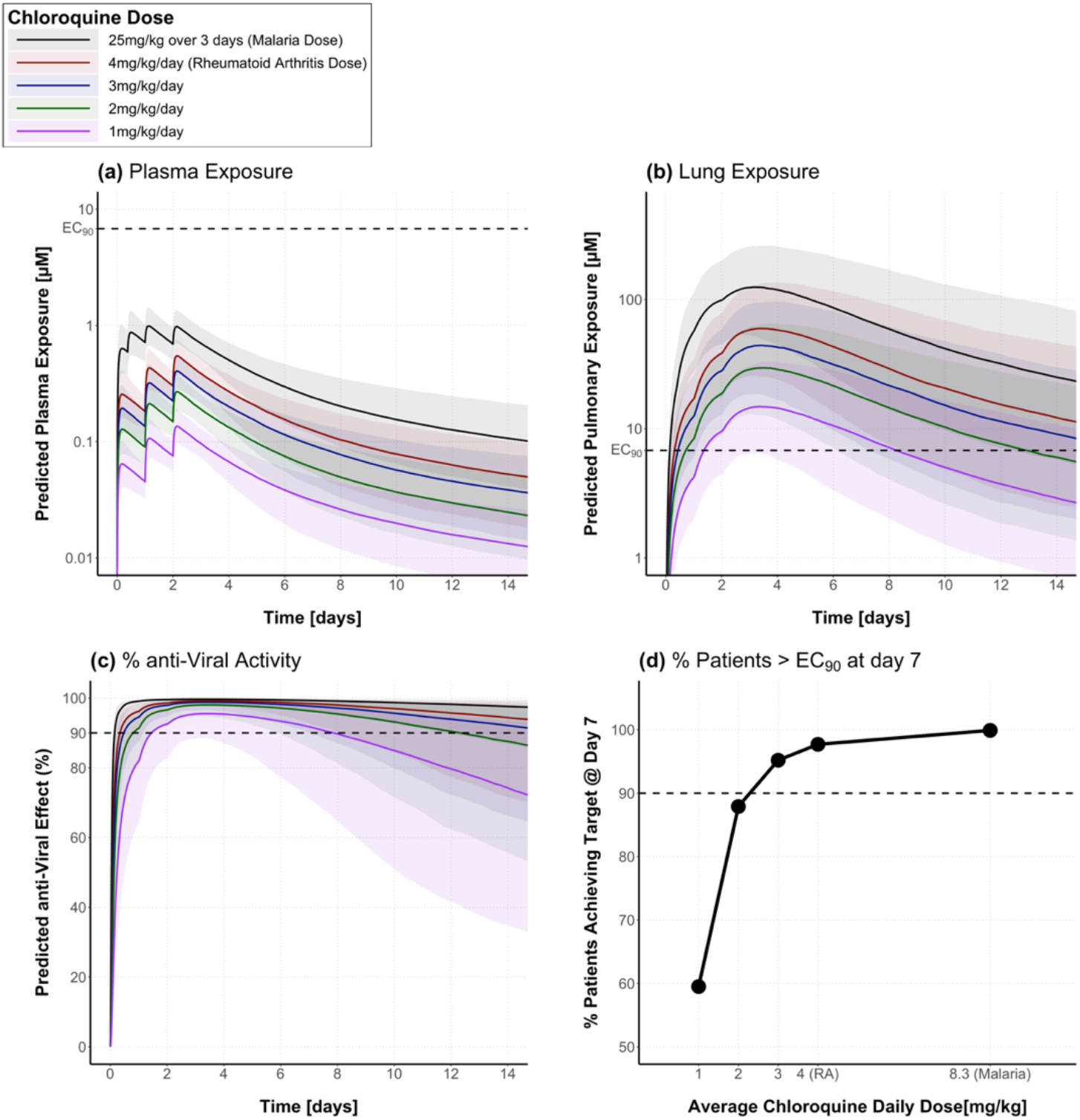
Predicted CQ Exposure in humans with a range of different dosing regimens. All doses are simulated as once daily for three days except the malaria dose which is administered as 10mg/kg at time 0, 5mg/kg 8 hours later, and then 5mg/kg on the 24^th^ and 48^th^ hour after treatment initiation. (a) shows predicted total exposure in plasma, (b) in lung, (c) % anti-viral activity based on pulmonary exposure and (d) shows percentage of patients who will achieve pulmonary exposure above the EC_90_ at day 7.

In contrast, plasma exposures even with the highest simulated dose of 25mg/kg over 3 days are very unlikely to reach to EC_90_ levels with a total plasma C_max_ (bound and unbound drug) that is some 7 fold lower than the EC_90_ against the virus (**Figure 4a**).

### iv. Predicting CQ’s prophylactic activity with >7day coverage

We explored the potential utility of CQ as a prophylactic and for coverage longer than 7 days, as previously described. Data demonstrate that at least 5-day dosing regimens with daily doses ≤ 10mg/kg would be required for the majority (>90%) of the population to exceed the EC_90_ at the level of the lung over a 28-day period. Shorter dosing regimens (3 days) would be sufficient to maintain local pulmonary concentrations higher than the EC_50_ but not the EC_90_ for the same duration (**Figure 5a, c**). Once weekly regimens, however, afford longer cover with doses lesser than 7mg/kg/week that would allow a 28-day cover above the EC_90_. Only 3mg/kg/week dose is required to maintain CQ pulmonary exposure above the EC_50_ over the same period of 28 days (**Figure 5c**). We also explored the potential of a single dose to give covers for periods of 1, 2 or 4 weeks. A single dose of less than 10mg/kg is unlikely to achieve pulmonary exposures above the EC_90_ for longer than 7 days. A single dose of 10mg/kg, however, can achieve a cover above the EC_50_ for 28 days (**Figure 5c**). At the plasma level however, none of the simulated dosing regimens can achieve any significant coverage over the EC_50_ or EC_90_ (**Figure 4a**).

**Figure 5.**
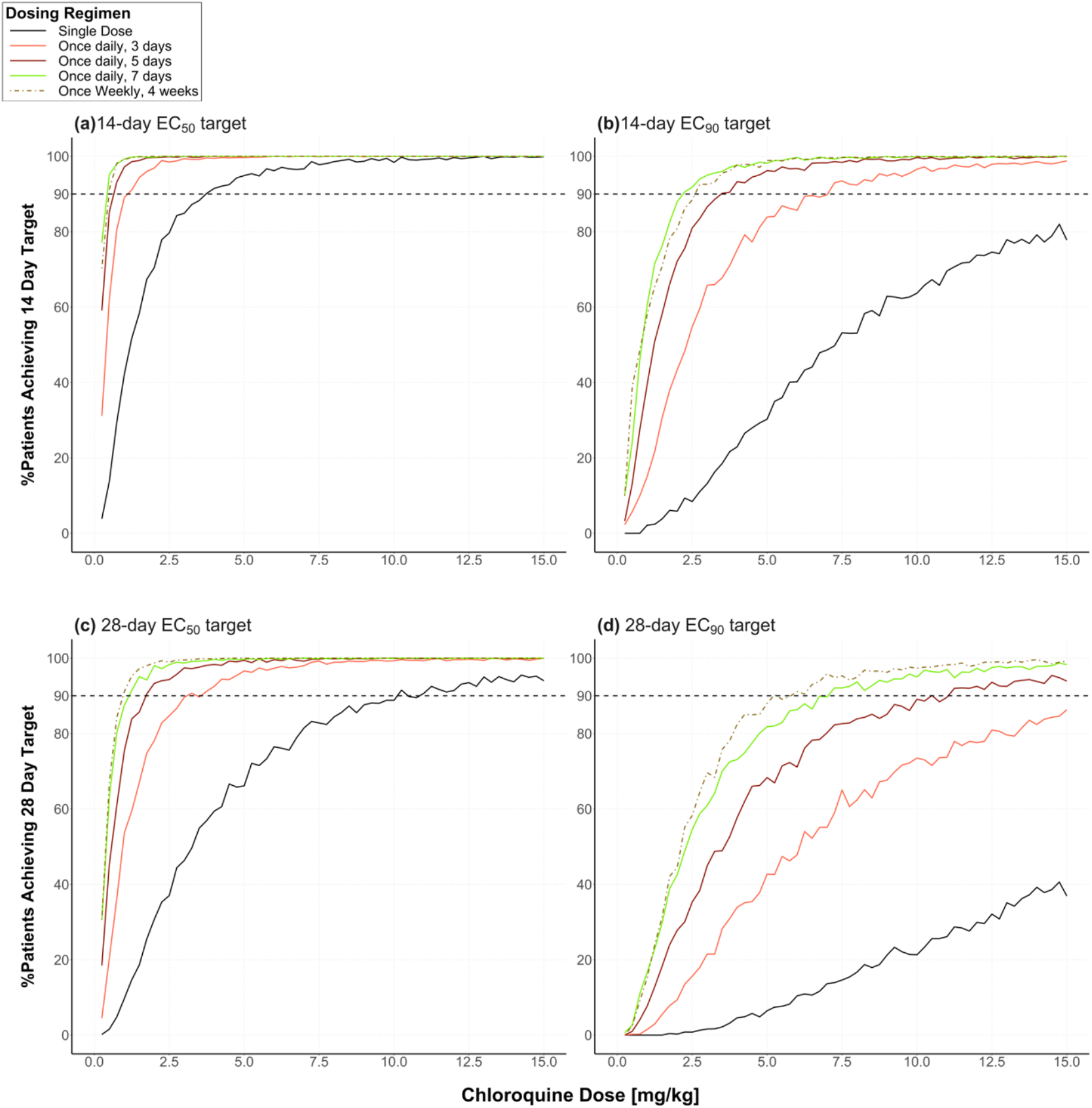
Monte Carlo simulations showing the relationship between administered CQ dose and percentage of patients achieving the target of pulmonary concentrations >EC_50_ (a, c) or EC_90_ (b, d) against SARS-CoV-2 for >90% of a 14-day (a, b) or 28-day (c, d) duration. Different lines represent different dosing regimens ranging from single dose (black line) to daily doses for up to 7 days (green line) or a once weekly dose (dashed brown line).

## DISCUSSION

Although controlled clinical trials are planned, there is little data available concerning CQ’s clinical efficacy against SARS-CoV2. Using previously published data it is possible to predict the lung exposure of CQ and HCQ which may be modelled in the context of the anti-viral activity against SARS-CoV2 (31). In this work we focus on CQ, where we use a population PK approach for dose selection that considers PK variation aided by using a combined pharmacodynamic dimension that estimates the real-time overall efficacy of the drug.

Although CQ is generally considered to be a safe drug, its use is not entirely risk-free and the drug is known to have a narrow therapeutic index, particularly as the dose increases marginally above that used for malaria treatment (32). The goal of this study was to establish the absolute minimum safe dose(s) that are required to deliver efficacious anti-viral activity, with the major assumption that the main driver for therapy is pulmonary drug exposure and not systemic exposure. Data available for the clinical selection of doses is dependent on regimens used for other diseases, such as malaria (25mg/kg over 3 days) or RA (4mg/kg chronic doses). COVID-19 is completely distinct from other CQ indications, due to the involvement of lung tissue in viral replication and disease pathology – a very different scenario to that observed for malarial parasites is in place; for example the drug has >1µM EC_50_ against the SARAS-CoV-2 but <0.01µM IC_50_ against non-resistant malaria parasites (33).

Our analyses confirm the concern that at safe CQ doses, such as those used in malaria, CQ will not achieve therapeutically relevant concentrations if systemic exposure is the primary driver (**Figure 4a**). In contrast, if pulmonary exposure is the driver, therapeutic success would be expected. It can even be argued that doses as low as 3mg/kg for 3 days or even a single dose treatment ≥ 10mg/kg should achieve therapeutically efficacious concentrations within the lung for 7 days or more. Also, from a PK-PD perspective, the modelling suggests that splitting the dose can significantly reduce pulmonary C_max_ while maintaining efficacious exposure which in turn could reduce the risk of adverse events in man. A 3-day regimen with a 3mg/kg dose, mirrors the duration of the accepted anti-malarial dosing regimen and demonstrates that it provides a 7-day cover against SARS-CoV2 (assuming pulmonary exposure based PKPD). Interestingly, 14-day cover could be achieved with a 5-day regimen of the same dose or a 3-day regimen with a 5mg/kg dose (**Figure 5a, b**), again using pulmonary exposure as the driver. The ongoing trials will formally establish if it is systemic or pulmonary exposure that is the main driver of activity.

The simulations in this work did not take into account protein binding. This was considered irrelevant as the total plasma CQ concentrations (bound+unbound) were significantly lower than the concentrations required for antiviral activity (>7 fold less) and are likely to be further lowered when correcting for plasma protein binding (CQ plasma protein fraction unbound (f_u_) = 0.54 (34)). For lung simulations, a correction for protein binding was not necessary as the epithelial lung fluid (ELF) albumin level is approximately 0.5mg/dL (35), which is similar to the albumin level in the *in-vitro* assays containing 10% Foetal Bovine Serum (FBS). PK-PD simulations relating total lung concentration to activity were previously successful in predicting the clinical activity of various anti-tubercular drugs without the need to correct for protein binding within the lung as the protein content was assumed to be similar here (36-38).

We also explored the potential of using CQ as a prophylactic treatment. Using the same modelling approach, we show that maintaining long duration pulmonary coverage in large populations is unlikely to be achieved using a single dose no matter what the driver of efficacy is. Instead, if pulmonary exposure is key, an optimal dosing regimen of once weekly at a dose of no more than 10mg/kg can achieve adequate cover whilst reducing the risk of undesirable side effects, associated with the chronic dosing of this drug. Simulations also indicate that, after the initial 3-days of treatment, weekly, not daily doses, may be administered to ensure that efficacious pulmonary levels of CQ are maintained.

This work addresses a priority concern arising from efforts to find repurposing opportunities for COVID-19. Specifically, can the *in-vitro* anti-SARS-CoV-2 potential of CQ be translated into human efficacy at doses that are safe? Our data suggests that if it is systemic exposure that needs to exceed *in-vitro* EC_90_s, CQ will not demonstrate efficacy at doses that are safe in man. In contrast pulmonary CQ concentrations that exceed these EC_90_s are readily achieved at moderate CQ dosages. In addition to the caveats described, it is important to also note that the assumption that the predicted pulmonary drug concentrations reflect drug concentrations at the local site of viral invasion and replication which may not be correct. Our modelling framework will be useful in defining optimised dosage regimens once ongoing clinical trials begin to readout. If these trials demonstrate efficacy it would suggest that lung rather than blood drug concentrations drive efficacy. Under these circumstances it may be possible to reduce CQ dosage and durations for this priority disease indication which in turn will deliver benefits in terms of drug supply and safety for a COVID-19 treatment/prophylactic with the potential for rapid global deployment.

## STUDY HIGHLIGHTS

- **What is the current knowledge on the topic?** CQ has been shown to exhibit *in-vitro* anti-viral activity against SARS-CoV-2 and is being trialled in patients with COVID-19.
- **What question did this study address?** Given CQ’s demonstrated *in-vitro* activity against SARS-CoV-2, does the drug achieve systemic and/or lung exposure levels that would allow for the clinical translation of this activity in man?
- **What does this study add to our knowledge?** CQ is unlikely to achieve systemic exposures that would exert any significant anti-viral activity given its *in-vitro* pharmacodynamic profile. However, its pulmonary exposure is high enough to allow for safe dosing levels that would achieve significantly efficacious pulmonary concentrations.
- **How might this change clinical pharmacology or translational science?** The study will help elucidate whether the main driver of activity of CQ in man is systemic or lung exposure depending on the outcome of ongoing clinical trials. If lung exposure is the driver for CQ’s activity, the simulations here show that currently suggested dosages can be significantly lowered for therapy or prophylaxis.

## Data Availability

Any required data will be made available upon contact.

## AUTHOR CONTRIBUTIONS

**GA, RR & HP** contributed to Manuscript writing, designing and performing research and analysing data. **SP, DH, PO, AO, SA and GB** contributed to Manuscript writing, and designing research.

## ACKNOWLEDGEMENTS

This work was supported in part by the Medical Research Council (MR/S00467X/1 G.A.B. and S.A.W.). G.A. acknowledges funding from the MRC Skills Development Fellowship.

## REFERENCES

(1) Adhanom, T. WHO Director-General’s opening remarks at the media briefing on COVID-19 - 11 March 2020. (2020).

(2) Dong, E., Du, H. & Gardner, L. An interactive web-based dashboard to track COVID-19 in real time. Lancet Infect Dis, (2020).

(3) Wang, M. et al. Remdesivir and chloroquine effectively inhibit the recently emerged novel coronavirus (2019-nCoV) in vitro. Cell Res 30, 269–71 (2020).

(4) Gao, J., Tian, Z. & Yang, X. Breakthrough: Chloroquine phosphate has shown apparent efficacy in treatment of COVID-19 associated pneumonia in clinical studies. Biosci Trends 14, 72–3 (2020).

(5) Rome, B.N. & Avorn, J. Drug Evaluation during the Covid-19 Pandemic. N Engl J Med, (2020).

(6) ClinicalTrials.gov [Internet]. Bethesda (MD): National Library of Medicine (US). 2000 Feb 29. Identifier NCT04303507, Chloroquine/ Hydroxychloroquine Prevention of Coronavirus Disease (COVID-19) in the Healthcare Setting (COPCOV); 2020 Mar 11 Available from: https://clinicaltrials.gov/ct2/show/NCT04303507.

(7) Keyaerts, E., Vijgen, L., Maes, P., Neyts, J. & Van Ranst, M. In vitro inhibition of severe acute respiratory syndrome coronavirus by chloroquine. Biochem Biophys Res Commun 323, 264–8 (2004).

(8) ACTIVITY of a new antimalarial agent, chloroquine (SN 7618). J Am Med Assoc 130, 1069 (1946).

(9) Ladeia-Andrade, S. et al. Monitoring the Efficacy of Chloroquine-Primaquine Therapy for Uncomplicated Plasmodium vivax Malaria in the Main Transmission Hot Spot of Brazil. Antimicrob Agents Chemother 63, (2019).

(10) Blignaut, M., Espach, Y., van Vuuren, M., Dhanabalan, K. & Huisamen, B. Revisiting the Cardiotoxic Effect of Chloroquine. Cardiovasc Drugs Ther 33, 1–11 (2019).

(11) Borba, M.G.S. et al. Effect of High vs Low Doses of Chloroquine Diphosphate as Adjunctive Therapy for Patients Hospitalized With Severe Acute Respiratory Syndrome Coronavirus 2 (SARS-CoV-2) Infection: A Randomized Clinical Trial. JAMA Network Open 3, e208857–e (2020).

(12) Megarbane, B. Chloroquine and hydroxychloroquine to treat COVID-19: between hope and caution. Clin Toxicol (Phila), 1–2 (2020).

(13) Wu, Z. & McGoogan, J.M. Characteristics of and Important Lessons From the Coronavirus Disease 2019 (COVID-19) Outbreak in China: Summary of a Report of 72314 Cases From the Chinese Center for Disease Control and Prevention. JAMA, (2020).

(14) Stokkermans, T.J. & Trichonas, G. Chloroquine And Hydroxychloroquine Toxicity. In: StatPearls (Treasure Island (FL), 2020).

(15) WHO. (1995). WHO Model Prescribing Information: Drugs Used in Parasitic Diseases - Second Edition (Geneva, Switzerland, 1995).

(16) Mackenzie, A.H. Dose refinements in long-term therapy of rheumatoid arthritis with antimalarials. Am J Med 75, 40–5 (1983).

(17) Nika, M., Blachley, T.S., Edwards, P., Lee, P.P. & Stein, J.D. Regular examinations for toxic maculopathy in long-term chloroquine or hydroxychloroquine users. JAMA Ophthalmol 132, 1199–208 (2014).

(18) Karunajeewa, H.A. et al. Pharmacokinetics of chloroquine and monodesethylchloroquine in pregnancy. Antimicrob Agents Chemother 54, 1186–92 (2010).

(19) Adelusi, S.A. & Salako, L.A. Tissue and blood concentrations of chloroquine following chronic administration in the rat. J Pharm Pharmacol 34, 733–5 (1982).

(20) R Core Team (2019). R: A language and environment for statistical computing. R Foundation for Statistical Computing, Vienna, Austria. URL https://www.R-project.org/.

(21) Monolix version 2019R2. Antony, France: Lixoft SAS, 2019. http://lixoft.com/products/monolix/.

(22) Adelusi, S.A. & Salako, L.A. Kinetics of the distribution and elimination of chloroquine in the rat. General Pharmacology: The Vascular System 13, 433–7 (1982).

(23) Prevention, C.f.D.C.a. Anthropometric Reference Data for Children and Adults: United States, 2011–2014. <https://www.cdc.gov/nchs/data/series/sr_03/sr03_039.pdf> (2016). Accessed 17/10/2019 2019.

(24) Bosgra, S., Eijkeren, J.v., Bos, P., Zeilmaker, M. & Slob, W. An improved model to predict physiologically based model parameters and their inter-individual variability from anthropometry. Crit Rev Toxicol 42, 751–67 (2012).

(25) NSCEP. (1994). Physiological Paramenter Values for PBPK Models 142 (1994).

(26) Peters, S. Evaluation of a Generic Physiologically Based Pharmacokinetic Model for Lineshape Analysis. Clin Pharmacokinet 47, 261–75 (2008).

(27) Rodgers, T., Leahy, D. & Rowland, M. Physiologically Based Pharmacokinetic Modeling 1: Predicting the Tissue Distribution of Moderate-to-Strong Bases. Journal of Pharmaceutical Sciences 94, 1259–76 (2005).

(28) Sager, J.E., Yu, J., Ragueneau-Majlessi, I. & Isoherranen, N. Physiologically Based Pharmacokinetic (PBPK) Modeling and Simulation Approaches: A Systematic Review of Published Models, Applications, and Model Verification. Drug Metab Dispos 43, 1823–37 (2015).

(29) Gustafsson, L.L. et al. Disposition of chloroquine in man after single intravenous and oral doses. Br J Clin Pharmacol 15, 471–9 (1983).

(30) Ono, C., Yamada, M. & Tanaka, M. Absorption, distribution and excretion of 14C-chloroquine after single oral administration in albino and pigmented rats: binding characteristics of chloroquine-related radioactivity to melanin in-vivo. J Pharm Pharmacol 55, 1647–54 (2003).

(31) Yao, X. et al. In Vitro Antiviral Activity and Projection of Optimized Dosing Design of Hydroxychloroquine for the Treatment of Severe Acute Respiratory Syndrome Coronavirus 2 (SARS-CoV-2). Clin Infect Dis, (2020).

(32) Davis, T.M., Syed, D.A., Ilett, K.F. & Barrett, P.H. Toxicity related to chloroquine treatment of resistant vivax malaria. Ann Pharmacother 37, 526–9 (2003).

(33) Srivastava, K., Agarwal, P., Soni, A. & Puri, S.K. Correlation between in vitro and in vivo antimalarial activity of compounds using CQ-sensitive and CQ-resistant strains of Plasmodium falciparum and CQ-resistant strain of P. yoelii. Parasitol Res 116, 1849–54 (2017).

(34) Charman, S.A. et al. An in vitro toolbox to accelerate anti-malarial drug discovery and development. Malar J 19, 1 (2020).

(35) Roberts, C.M. et al. Changes in epithelial lining fluid albumin associated with smoking and interstitial lung disease. Eur Respir J 6, 110–5 (1993).

(36) Donnellan, S. et al. Intracellular Pharmacodynamic Modeling Is Predictive of the Clinical Activity of Fluoroquinolones against Tuberculosis. Antimicrob Agents Chemother 64, (2019).

(37) Aljayyoussi, G., Donnellan, S., Ward, S.A. & Biagini, G.A. Intracellular PD Modelling (PDi) for the Prediction of Clinical Activity of Increased Rifampicin Dosing. Pharmaceutics 11, (2019).

(38) Aljayyoussi, G. et al. Pharmacokinetic-Pharmacodynamic modelling of intracellular Mycobacterium tuberculosis growth and kill rates is predictive of clinical treatment duration. Sci Rep 7, 502 (2017).

